# Association of Bacille Calmette-Guérin (BCG), Adult Pneumococcal and Adult Seasonal Influenza Vaccines with Covid-19 Adjusted Mortality Rates in Level 4 European countries

**DOI:** 10.1101/2020.06.03.20121624

**Authors:** Joe Gallagher, Chris Watson, Mark Ledwidge

## Abstract

**Introduction:** Non-specific effects of vaccines have gained increasing interest during the Covid-19 pandemic. In particular, population use of BCG vaccine has been associated with improved outcomes. This study sought to determine the association of population use of BCG, adult pneumococcal and adult seasonal influenza vaccination with Covid-19 mortality when adjusted for a number of confounding variables.

**Methods:** Using publicly available data, mortality adjusted for the timeframe of crisis, population size and population characteristics was calculated. The primary analysis was the relationship between each of the day 15 and day 30 standardised mortality rates and BCG, adult pneumococcal and influenza vaccination scores using unadjusted measures and with adjustment for population structure and case fatality rates. Secondary analyses were measures of case increases and mortality increases from day 15 to day 30 for each of the relative vaccination scores. Finally, we also analysed the peak Z score reflecting increases in total mortality from historical averages reported by EuroMOMO (Euromomo.eu),

**Results:** Following adjustment for the effects of population size, median age, population density, the proportion of population living in an urban setting, life-expectancy, the elderly dependency ratio (or proportion over 65 years), net migration, days from day 1 to lockdown and case-fatality rate, only BCG vaccination score remained significantly associated with Covid-19 mortality at day 30. In the best fit model, BCG vaccination score was associated with a 64% reduction in log(10) mortality per 10 million population (OR 0.362 reduction [95% CI 0.188 to 0.698]), following adjustment for population size, median age, density, urbanization, elderly dependency ratio, days to lockdown, yearly migration and case fatality rate.

**Conclusion:** BCG vaccine was associated with reduced mortality rates in level 4 countries while adult pneumococcal and adult seasonal influenza vaccines were not when adjusted for a number of confounding variables. A number of trials are ongoing to determine if BCG is protective against severe Covid-19 infection.

## Introduction

Non-specific, “priming” effects of vaccines have rapidly gained interest recently during the Covid 19 pandemic. While Bacille Calmette-Guérin (BCG), pneumococcal, influenza and others vaccines have been associated with beneficial effects beyond those targeting the illness they are designed to prevent(1–5), considerable public interest has followed ecological studies suggesting that countries with universal BCG vaccination policies have lower mortality with Covid 19(6–9).

However, the studies suggesting beneficial effects of the BCG vaccine to date are limited in at least 4 important ways. First, countries have very specific timeframes in the Covid 19 outbreak, yet none of the mortality comparisons to date adjust for timeframe as well as population size. Second, the population structure differs considerably in the countries studied, resulting in confounding by factors such as population size, population density, urbanisation and elderly dependency ratio. Thirdly, the apparent case fatality rate is very different in study countries and, in addition to population structure factors mentioned above, is dependent on the case definition, testing strategy and availability of SARS-CoV-2testing, which has been different in the countries. Finally, documentation of Covid19 deaths in level 1–3 (Low and Middle Income) countries is limited because a high proportion of deaths occur at home without medical assistance and my not be accounted for properly in government records. Indeed, these concerns have led to the WHO issuing a scientific brief highlighting that is no evidence currently that the BCG vaccine protects people against infection with COVID-19 virus and that further study is required(10).

Accordingly, while the results of prospective studies are awaited, we investigated the association between adult influenza, pneumococcal and BCG vaccination rates and standardised Covid 19 mortality outcomes in 25 level 4 European countries, taking into account the important confounders described above.

## Methods

Using publicly available mortality data from the John’s Hopkins Coronavirus Research Centre and demographic information presented on *http://worldometers.info*, we constructed curves of cumulative mortality adjusted for timeframe of crisis and population size, as previously described in a rapid response to an editorial on SARS-CoV-2testing and comparative international mortality rates[11a]. In order to standardise the timeframe, we designated the baseline (day 1) as the day that mortality rates exceeded 1 per 5 million population. We calculated the day 15 and day 30 mortality rate per 10 million population for each of the following 25 level 4 European countries Austria, Belgium, Czechia, Denmark, Estonia, Finland, France, Germany, Greece, Hungary, Iceland, Ireland, Italy, Lithuania, Luxembourg, Netherlands, Norway, Poland, Portugal, Slovakia, Slovenia, Spain, Sweden, Switzerland, United Kingdom. We also obtained recent data on population structure including population size, the median age (years), population density (persons/Km2), the proportion of population living in an urban setting, life-expectancy (years), the elderly dependency ratio (proportion of people over 65 years divided by the proportion aged 15–65) and net migration (expressed per 10 million population). In multivariable modelling we also included the case fatality rate on day 21 to account for differences in testing rates, testing strategy and case definition as well as the relative number of days between baseline (day 1) and lockdown.

In order to standardise the estimation of the proportion of the population covered with BCG vaccination, we generated a BCG vaccination score by dividing the median life expectancy by the total number of years that BCG vaccination was available and recommended for all children. We adjusted this calculation by multiplying with a measure of vaccination programme efficacy, using the most recent vaccination coverage rate for either BCG or universal childhood vaccination rates as appropriate. In the case of two countries that do not routinely recommend BCG vaccination and where few details were available (Iceland, Luxembourg) we used referenced estimates of the total population covered by BCG vaccination. Similarly, for pneumococcal vaccination, we generated a pneumococcal vaccination score by estimating the proportion of older people covered by pneumococcal vaccination over the past decade using a variety of referenced sources in circumstances where uptake is considered relatively low in most European countries, despite recommendations for their use in over 65s and vulnerable populations. Finally, we created an influenza vaccination score as the proportion of older adults with seasonal influenza vaccination from most recent sources provided by the European Centre for Disease Control and other referenced reports.

The primary analysis of our report was the relationship between each of the day 15 and day 30 standardised mortality rates and BCG, pneumococcal and Influenza vaccination scores using unadjusted measures and with adjustment for population structure and case fatality rates. Secondary analyses were measures of case increases and mortality increases from day 15 to day 30 for each of the relative vaccination scores. Finally, we also analysed the peak Z score reflecting increases in total mortality from historical averages reported by EuroMOMO (Euromomo.eu), a European mortality monitoring activity, aiming to detect and measure excess deaths related to seasonal influenza, pandemics and other public health threats, supported by the European Centre for Disease Prevention and Control (ECDC) and the World Health Organization (WHO).

Primary and secondary outcomes were analysed using generalized linear modeling with a Poisson outcome distribution for adjusted mortality rates. Analyses were performed both with and without adjustment for the effects of population size, the median age, population density, the proportion of population living in an urban setting, life-expectancy, the elderly dependency ratio (or proportion over 65 years), net migration, days from day 1 to lockdown and case-fatality rate. Variables such as day 15 and day 30 mortality rates, case fatality rates, population size, population density and life expectancy, were not normally distributed across the countries and were log-transformed before inclusion in the generalized linear model. Nominal statistical significance was set at P< 0.05 with Bonferroni correction for the 3 pairwise correlations involved in the primary analyses (three vaccination scores linked to day 30 mortality rates), meaning a P value of 0.05/3=0.0167 was considered statistically significant. Descriptive data are presented as either mean ± SD or median (25th: 75th percentile) for normally and non-normally distributed continuous variables, respectively. Frequencies and percentages (in parentheses) summarize categorical variables. Graphs were presented using GraphPad Prism v6 and statistical analysis was performed using the R statistical software package (Version 3.6.2, 2019–12–12, Copyright 2019 The R Foundation).

## Results

The population structure of the 25 countries are presented in Table 1.

**TABLE 1.** Population structure of the 25 countries included in the analysis, obtained from http://worldometers.info, April 2020. Abbreviation: UK, United Kingdom.

| Country | Population | Median age (years) | Over 65 years (%) | Life expectancy (years) | Population density (P/Km<sup>2</sup>) | Urban population proportion | Elderly Dependency Ratio | Yearly migration (%) |
| --- | --- | --- | --- | --- | --- | --- | --- | --- |
| Austria | 9006398 | 43.5 | 19.0 | 82.1 | 109 | 0.57 | 0.31 | 0.0057 |
| Belgium | 11589623 | 41.9 | 18.8 | 82.2 | 383 | 0.98 | 0.31 | 0.0044 |
| Czechia | 10708981 | 43.2 | 19.4 | 79.9 | 139 | 0.74 | 0.29 | 0.0018 |
| Denmark | 5792202 | 42.3 | 19.8 | 81.4 | 137 | 0.88 | 0.33 | 0.0035 |
| Estonia | 1326535 | 42.4 | 19.6 | 79.2 | 31 | 0.68 | 0.31 | 0.0007 |
| Finland | 5540720 | 43.1 | 21.7 | 82.5 | 18 | 0.86 | 0.35 | 0.0015 |
| France | 65273511 | 42.3 | 20.3 | 83.1 | 119 | 0.82 | 0.33 | 0.0022 |
| Germany | 83783942 | 45.7 | 21.5 | 81.9 | 240 | 0.76 | 0.35 | 0.0032 |
| Greece | 10423054 | 45.6 | 21.7 | 82.8 | 81 | 0.85 | 0.33 | -0.0048 |
| Hungary | 9660351 | 43.3 | 19.2 | 77.3 | 107 | 0.72 | 0.28 | -0.0025 |
| Iceland | 341243 | 37.5 | 14.8 | 83.5 | 3 | 0.94 | 0.23 | 0.0065 |
| Ireland | 4937786 | 38.2 | 13.9 | 82.8 | 72 | 0.63 | 0.22 | 0.0113 |
| Italy | 60461826 | 47.3 | 22.8 | 84.0 | 206 | 0.70 | 0.38 | -0.0015 |
| Lithuania | 2722289 | 45.1 | 19.7 | 76.4 | 43 | 0.71 | 0.29 | -0.0135 |
| Luxembourg | 625978 | 39.7 | 14.1 | 82.8 | 242 | 0.88 | 0.22 | 0.0166 |
| Netherlands | 17134872 | 43.3 | 18.8 | 82.8 | 508 | 0.93 | 0.30 | 0.0022 |
| Norway | 5421241 | 39.8 | 17.0 | 92.9 | 15 | 0.83 | 0.27 | 0.0079 |
| Poland | 37846611 | 41.7 | 17.5 | 79.3 | 124 | 0.60 | 0.24 | -0.0011 |
| Portugal | 10196709 | 46.2 | 22.0 | 82.7 | 111 | 0.67 | 0.35 | -0.0029 |
| Slovakia | 5459642 | 41.2 | 15.6 | 78.0 | 114 | 0.54 | 0.22 | 0.0005 |
| Slovenia | 2078938 | 44.5 | 19.6 | 81.9 | 103 | 0.55 | 0.29 | 0.0001 |
| Spain | 46754778 | 44.9 | 19.4 | 84.0 | 94 | 0.80 | 0.31 | 0.0004 |
| Sweden | 10099265 | 41.1 | 21.0 | 83.3 | 25 | 0.88 | 0.34 | 0.0063 |
| Switzerland | 8654622 | 43.1 | 18.6 | 84.3 | 219 | 0.74 | 0.29 | 0.0074 |
| United Kingdom | 67886011 | 40.5 | 18.4 | 81.8 | 281 | 0.83 | 0.31 | 0.0053 |
| **Median** | **9660351** | **43.1** | **19.4** | **82.5** | **111** | **0.76** | **0.31** | **0.0022** |
| *Quartile 1* | *5421241* | *41.2* | *18.4* | *81.4* | *72* | *0.68* | *0.28* | *0.0001* |
| *Quartile 3* | *17134872* | *44.5* | *20.3* | *83.1* | *206* | *0.86* | *0.33* | *0.0057* |

The date of exceeding 1 SARS-CoV-2death per 5 million population, date of lockdown and cases, deaths and case fatality rates (CFR) on days 15 and 30 are presented in Table 2.

**TABLE 2.** Outcome data for the 25 countries included in the analysis, including cases, deaths, case fatality rates (CFR) and changes in deaths and cases between day 15 and day 30. Data obtained from http://worldometers.info, April 2020. *Day 1 refers to the date that the country exceeded 1 death per 5 million population. **The Days to Lockdown refers to the relative number of days from Day 1 to date of lock-down. *** Refers to the change in deaths and cases per 10 million population between days 15 and 30. Abbreviations: CFR Case Fataility Rate; UK, United Kingdom.

| Country | Day 1* | Date of Lock-down | Days To Lock-down** | Cases / 10m day 15 | Cases / 10m day 30 | Deaths / 10m day 15 | Deaths / 10m day 30 | CFR day 15 | CFR day 30 | $\Delta$ Deaths / 10m day 15-30 *** | $\Delta$ Cases / 10m day 15-30 *** |
| --- | --- | --- | --- | --- | --- | --- | --- | --- | --- | --- | --- |
| Austria | 16/03/2020 | 16/03/2020 | 0 | 11303 | 16205 | 142.12 | 478.55 | 0.013 | 0.030 | 336.43 | 490.21 |
| Belgium | 11/03/2020 | 18/03/2020 | 7 | 5380 | 23009 | 189.82 | 2604.92 | 0.035 | 0.113 | 2415.09 | 1762.96 |
| Czechia | 24/03/2020 | 16/03/2020 | -8 | 4685 | 6711 | 82.17 | 196.10 | 0.018 | 0.029 | 113.92 | 202.63 |
| Denmark | 15/03/2020 | 11/03/2020 | -4 | 4449 | 11241 | 132.94 | 516.21 | 0.030 | 0.046 | 383.27 | 679.19 |
| Estonia | 25/03/2020 | 13/03/2020 | -12 | 8933 | 12099 | 180.92 | 346.77 | 0.020 | 0.029 | 165.85 | 316.61 |
| Finland | 25/03/2020 | 27/03/2020 | 2 | 4489 | 7932 | 72.19 | 319.45 | 0.016 | 0.040 | 247.26 | 344.36 |
| France | 07/03/2020 | 17/03/2020 | 10 | 2454 | 13857 | 103.26 | 1365.18 | 0.042 | 0.099 | 1261.92 | 1140.34 |
| Germany | 16/03/2020 | 23/03/2020 | 7 | 8571 | 16083 | 92.50 | 454.02 | 0.011 | 0.028 | 361.53 | 751.28 |
| Greece | 14/03/2020 | 23/03/2020 | 9 | 1109 | 2058 | 37.42 | 94.98 | 0.034 | 0.046 | 57.56 | 94.89 |
| Hungary | 20/03/2020 | 28/03/2020 | 8 | 645 | 1983 | 26.91 | 195.65 | 0.042 | 0.099 | 168.73 | 133.85 |
| Iceland | 21/03/2020 | 13/03/2020 | -8 | 41525 | 51957 | 117.22 | 293.05 | 0.003 | 0.006 | 175.83 | 1043.24 |
| Ireland | 11/03/2020 | 14/03/2020 | 3 | 3684 | 16382 | 38.48 | 581.23 | 0.010 | 0.035 | 542.75 | 1269.80 |
| Italy | 27/02/2020 | 09/03/2020 | 11 | 2921 | 14306 | 209.39 | 1510.71 | 0.072 | 0.108 | 1301.32 | 1138.54 |
| Lithuania | 20/03/2020 | 16/03/2020 | -4 | 2384 | 4768 | 33.06 | 128.57 | 0.014 | 0.027 | 95.51 | 238.40 |
| Luxembourg | 13/03/2020 | 18/03/2020 | 5 | 29250 | 52414 | 287.55 | 1054.35 | 0.010 | 0.020 | 766.80 | 2316.38 |
| Netherlands | 09/03/2020 | 16/03/2020 | 7 | 3245 | 11993 | 161.08 | 1311.94 | 0.050 | 0.109 | 1150.87 | 874.77 |
| Norway | 14/03/2020 | 12/03/2020 | -2 | 7902 | 12180 | 47.96 | 247.18 | 0.006 | 0.020 | 199.22 | 427.76 |
| Poland | 23/03/2020 | 13/03/2020 | -10 | 1166 | 2687 | 28.27 | 112.56 | 0.024 | 0.042 | 84.29 | 152.09 |
| Portugal | 19/03/2020 | 19/03/2020 | 0 | 9695 | 19305 | 241.25 | 673.75 | 0.025 | 0.035 | 432.49 | 961.00 |
| Slovakia | 06/04/2020 | 16/03/2020 | -21 | 2196 | 2577 | 25.64 | 49.45 | 0.012 | 0.018 | 23.81 | 38.10 |
| Slovenia | 14/03/2020 | 16/03/2020 | 2 | 3410 | 5830 | 52.91 | 678.23 | 0.016 | 0.045 | 625.32 | 241.95 |
| Spain | 07/03/2020 | 14/03/2020 | 7 | 6153 | 29232 | 379.00 | 2853.40 | 0.062 | 0.098 | 2474.40 | 2307.94 |
| Sweden | 15/03/2020 | NA | NA | 3988 | 11333 | 144.56 | 1022.85 | 0.036 | 0.090 | 878.28 | 734.41 |
| Switzerland | 08/03/2020 | 17/03/2020 | 9 | 10162 | 25712 | 138.65 | 948.63 | 0.014 | 0.037 | 809.97 | 1555.01 |
| UK | 14/03/2020 | 23/03/2020 | 9 | 2876 | 13054 | 245.85 | 1919.25 | 0.085 | 0.147 | 1673.39 | 1017.87 |
| **Median** | **15/03/2020** | **16/03/2020** | **2.5** | **4449** | **12180** | **117.22** | **516.21** | **0.020** | **0.040** | **383.27** | **734.41** |
| *Quartile 1* | *11/03/2020* | *13/03/2020* | *-4* | *2876* | *6711* | *47.96* | *247.18* | *0.013* | *0.029* | *168.73* | *241.95* |
| *Quartile 2* | *20/03/2020* | *18/03/2020* | *7.25* | *8571* | *16382* | *180.92* | *1054.35* | *0.036* | *0.098* | *878.28* | *1138.54* |

The BCG, pneumococcal and Influenza vaccination scores, datasources as well as the derivation values for BCG vaccination scores are presented in Table 3.

**TABLE 3.**
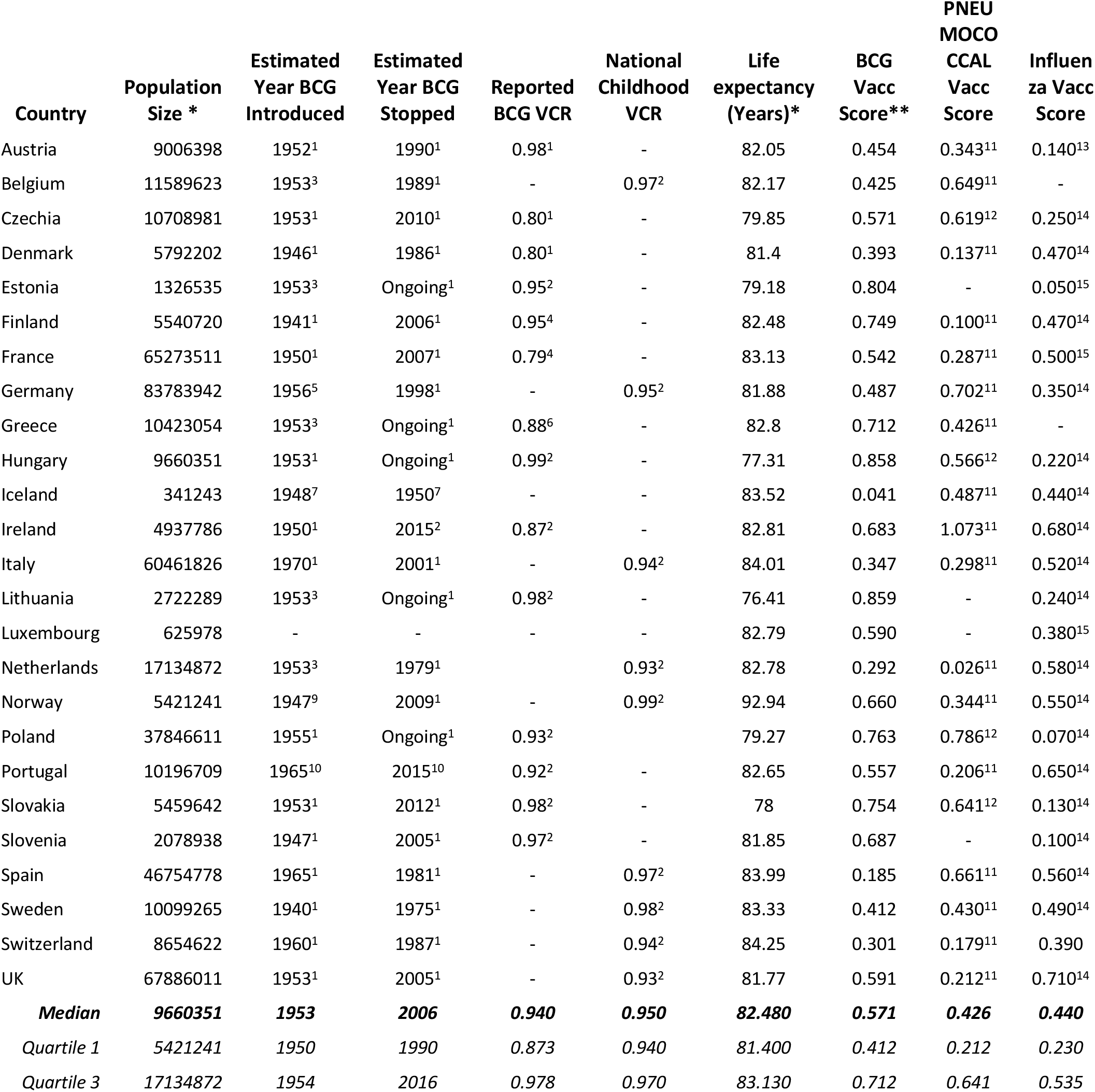
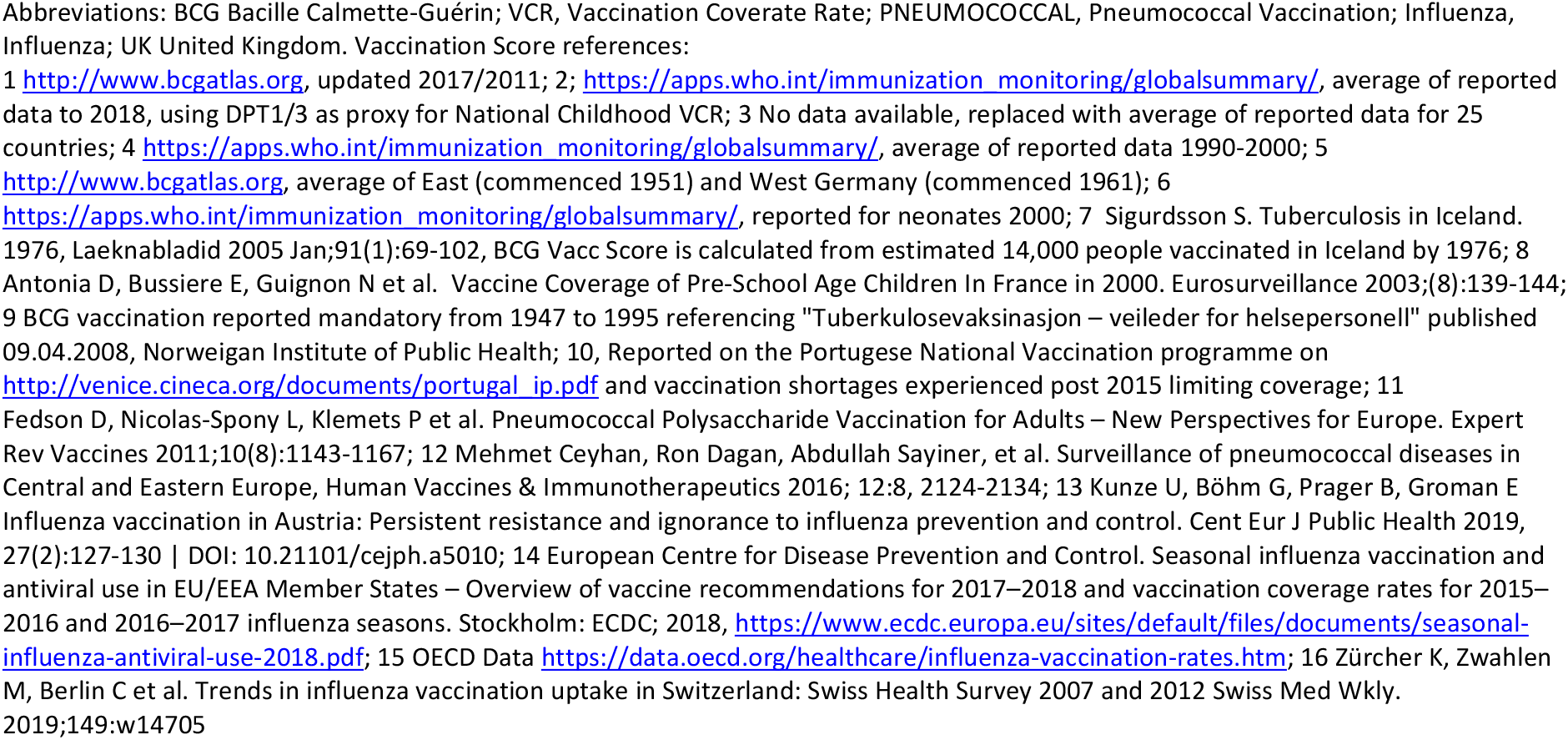
BCG, pneumococcal (PNEUMOCOCCAL) and influenza (Influenza) vaccination scores for the countries included in the analysis with referenced sources. *Population and Life Expectancy data were obtained from http://Worldometers.info, May 2020. **BCG Vacc Score was calculated by multiplying the estimated vaccination coverage rate (BCG if available or Dpt1/3 childhood vaccination coverage estimate if not) by the country’s average life expectancy divided by the number of years BCG vaccination availability.

| Country | Population Size * | Estimated Year BCG Introduced | Estimated Year BCG Stopped | Reported BCG VCR | National Childhood VCR | Life expectancy (Years)* | BCG Vacc Score** | PNEU MOCO CCAL Vacc Score | Influenza Vacc Score |
| --- | --- | --- | --- | --- | --- | --- | --- | --- | --- |
| Austria | 9006398 | 1952<sup>1</sup> | 1990<sup>1</sup> | 0.98<sup>1</sup> | - | 82.05 | 0.454 | 0.343<sup>11</sup> | 0.140<sup>13</sup> |
| Belgium | 11589623 | 1953<sup>3</sup> | 1989<sup>1</sup> | - | 0.97<sup>2</sup> | 82.17 | 0.425 | 0.649<sup>11</sup> | - |
| Czechia | 10708981 | 1953<sup>1</sup> | 2010<sup>1</sup> | 0.80<sup>1</sup> | - | 79.85 | 0.571 | 0.619<sup>12</sup> | 0.250<sup>14</sup> |
| Denmark | 5792202 | 1946<sup>1</sup> | 1986<sup>1</sup> | 0.80<sup>1</sup> | - | 81.4 | 0.393 | 0.137<sup>11</sup> | 0.470<sup>14</sup> |
| Estonia | 1326535 | 1953<sup>3</sup> | Ongoing<sup>1</sup> | 0.95<sup>2</sup> | - | 79.18 | 0.804 | - | 0.050<sup>15</sup> |
| Finland | 5540720 | 1941<sup>1</sup> | 2006<sup>1</sup> | 0.95<sup>4</sup> | - | 82.48 | 0.749 | 0.100<sup>11</sup> | 0.470<sup>14</sup> |
| France | 65273511 | 1950<sup>1</sup> | 2007<sup>1</sup> | 0.79<sup>4</sup> | - | 83.13 | 0.542 | 0.287<sup>11</sup> | 0.500<sup>15</sup> |
| Germany | 83783942 | 1956<sup>5</sup> | 1998<sup>1</sup> | - | 0.95<sup>2</sup> | 81.88 | 0.487 | 0.702<sup>11</sup> | 0.350<sup>14</sup> |
| Greece | 10423054 | 1953<sup>3</sup> | Ongoing<sup>1</sup> | 0.88<sup>6</sup> | - | 82.8 | 0.712 | 0.426<sup>11</sup> | - |
| Hungary | 9660351 | 1953<sup>1</sup> | Ongoing<sup>1</sup> | 0.99<sup>2</sup> | - | 77.31 | 0.858 | 0.566<sup>12</sup> | 0.220<sup>14</sup> |
| Iceland | 341243 | 1948<sup>7</sup> | 1950<sup>7</sup> | - | - | 83.52 | 0.041 | 0.487<sup>11</sup> | 0.440<sup>14</sup> |
| Ireland | 4937786 | 1950<sup>1</sup> | 2015<sup>2</sup> | 0.87<sup>2</sup> | - | 82.81 | 0.683 | 1.073<sup>11</sup> | 0.680<sup>14</sup> |
| Italy | 60461826 | 1970<sup>1</sup> | 2001<sup>1</sup> | - | 0.94<sup>2</sup> | 84.01 | 0.347 | 0.298<sup>11</sup> | 0.520<sup>14</sup> |
| Lithuania | 2722289 | 1953<sup>3</sup> | Ongoing<sup>1</sup> | 0.98<sup>2</sup> | - | 76.41 | 0.859 | - | 0.240<sup>14</sup> |
| Luxembourg | 625978 | - | - | - | - | 82.79 | 0.590 | - | 0.380<sup>15</sup> |
| Netherlands | 17134872 | 1953<sup>3</sup> | 1979<sup>1</sup> | - | 0.93<sup>2</sup> | 82.78 | 0.292 | 0.026<sup>11</sup> | 0.580<sup>14</sup> |
| Norway | 5421241 | 1947<sup>9</sup> | 2009<sup>1</sup> | - | 0.99<sup>2</sup> | 92.94 | 0.660 | 0.344<sup>11</sup> | 0.550<sup>14</sup> |
| Poland | 37846611 | 1955<sup>1</sup> | Ongoing<sup>1</sup> | 0.93<sup>2</sup> | - | 79.27 | 0.763 | 0.786<sup>12</sup> | 0.070<sup>14</sup> |
| Portugal | 10196709 | 1965<sup>10</sup> | 2015<sup>10</sup> | 0.92<sup>2</sup> | - | 82.65 | 0.557 | 0.206<sup>11</sup> | 0.650<sup>14</sup> |
| Slovakia | 5459642 | 1953<sup>1</sup> | 2012<sup>1</sup> | 0.98<sup>2</sup> | - | 78 | 0.754 | 0.641<sup>12</sup> | 0.130<sup>14</sup> |
| Slovenia | 2078938 | 1947<sup>1</sup> | 2005<sup>1</sup> | 0.97<sup>2</sup> | - | 81.85 | 0.687 | - | 0.100<sup>14</sup> |
| Spain | 46754778 | 1965<sup>1</sup> | 1981<sup>1</sup> | - | 0.97<sup>2</sup> | 83.99 | 0.185 | 0.661<sup>11</sup> | 0.560<sup>14</sup> |
| Sweden | 10099265 | 1940<sup>1</sup> | 1975<sup>1</sup> | - | 0.98<sup>2</sup> | 83.33 | 0.412 | 0.430<sup>11</sup> | 0.490<sup>14</sup> |
| Switzerland | 8654622 | 1960<sup>1</sup> | 1987<sup>1</sup> | - | 0.94<sup>2</sup> | 84.25 | 0.301 | 0.179<sup>11</sup> | 0.390 |
| UK | 67886011 | 1953<sup>1</sup> | 2005<sup>1</sup> | - | 0.93<sup>2</sup> | 81.77 | 0.591 | 0.212<sup>11</sup> | 0.710<sup>14</sup> |
| **Median** | **9660351** | **1953** | **2006** | **0.940** | **0.950** | **82.480** | **0.571** | **0.426** | **0.440** |
| *Quartile 1* | *5421241* | *1950* | *1990* | *0.873* | *0.940* | *81.400* | *0.412* | *0.212* | *0.230* |
| *Quartile 3* | *17134872* | *1954* | *2016* | *0.978* | *0.970* | *83.130* | *0.712* | *0.641* | *0.535* |

Abbreviations: BCG Bacille Calmette-Guérin; VCR, Vaccination Coverate Rate; PNEUMOCOCCAL, Pneumococcal Vaccination; Influenza, Influenza; UK United Kingdom. Vaccination Score references:

**FIGURE 1.**
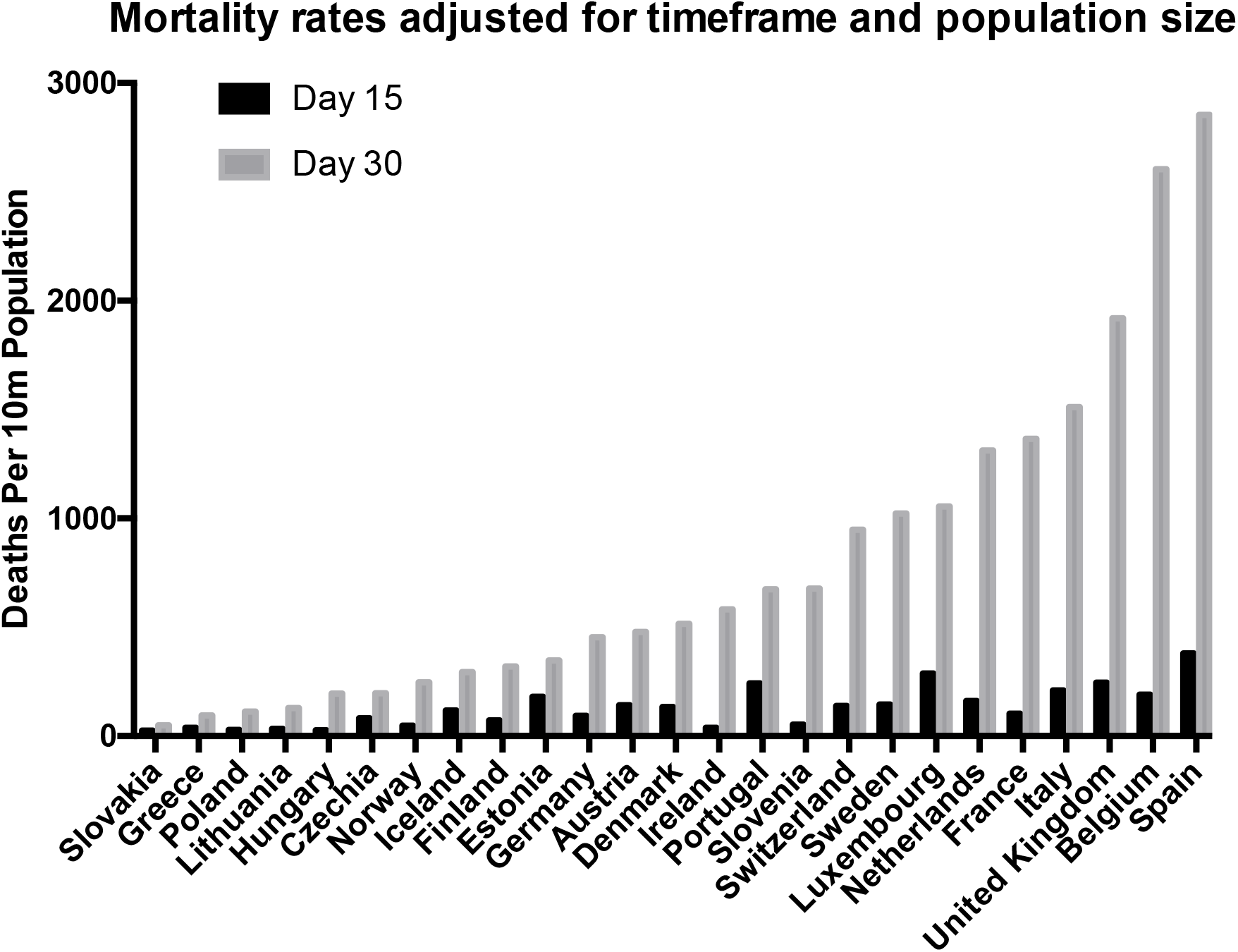
Day 15 and day 30 mortality per 10m population for the 25 countries included in the analysis, sorted in ascending order of day 30 mortality per 10m population. Data obtained from http://worldometers.info, April 2020

The unadjusted linear regression between log of deaths per 10 million on day 30 versus BCG, pneumococcal and influenza vaccination scores are presented in Figure 2A-C. Following adjustment for the effects of population size, median age, population density, the proportion of population living in an urban setting, life-expectancy, the elderly dependency ratio (or proportion over 65 years), net migration, days from day 1 to lockdown and case-fatality rate, BCG vaccination score remained significantly associated with Covid 19 mortality at day 30. In the best fit model, BCG vaccination score was associated with a 64% reduction in log(10) mortality per 10 million population (OR 0.362 reduction [95% CI 0.188 to 0.698]), following adjustment for population size, median age, density, urbanization, elderly dependency ratio, days to lockdown, yearly migration and case fatality rate. The log of case fatality rate (OR 2.526 [95% CI 1.147 to 5.563]) was the only other covariate that was significantly associated with Covid 19 mortality at day 30. There was no adjusted association between reported pneumococcal and influenza vaccination rates in older adults and Covid 19 mortality.

**FIGURE 2A-C.**
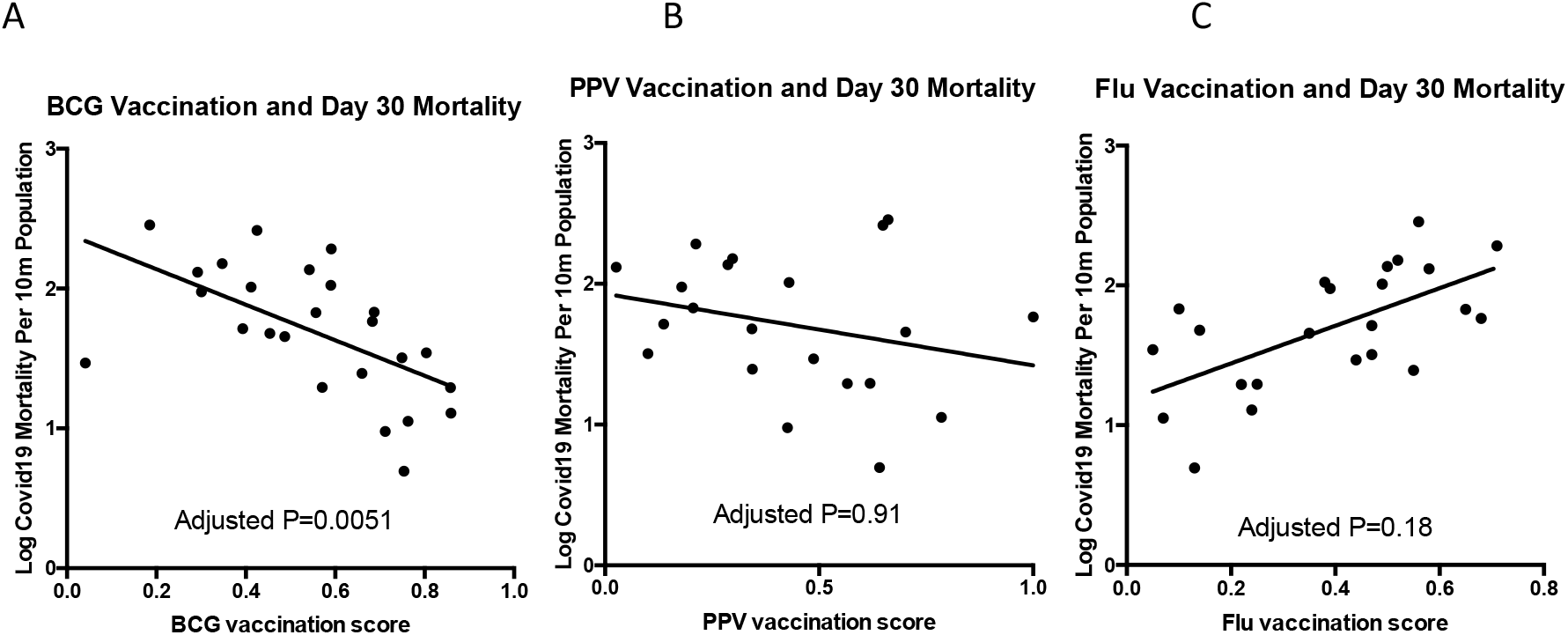
Primary outcome data for the correlation between cumulative Covid 19 mortality per ten million population at day 30 and the following: BCG vaccination score (A), unadjusted P = 0.0022; PNEUMOCOCCAL vaccination score (B), unadjusted P = 0.24; Influenza vaccination score (C), unadjusted P = 0.001. Following adjustment for the effects of population size, median age, population density, the proportion of population living in an urban setting, life-expectancy, the elderly dependency ratio (or proportion over 65 years), net migration, days from day 1 to lockdown and case-fatality rate, only BCG vaccination score remained significantly associated with Covid 19 mortality at day 30. Abbreviations: BCG, Bacille Calmette-Guérin; PNEUMOCOCCAL, pneumococcal; Influenza, influenza.

Similarly, as shown in Figure 3A-C, BCG vaccination score was significantly associated with day 15 log transformed mortality per 10 million population (HR 0.511 reduction [95% CI 0.277 to 0.943]), increase in cases between day 15 and day 30 (HR 0.398 reduction [95% CI 0.167 to 0.947]) and increase in mortality between day 15 and day 30 (HR 0.398 reduction [95% CI 0.167 to 0.947]). None of the other vaccination scores (PNEUMOCOCCAL or Influenza) were associated with these secondary outcomes in adjusted models. Finally, amongst those countries contributing total mortality data to the European Mortality Monitoring Centre, the relationship between BCG vaccination and peak Z-score remains significant, again with and without covariate adjustment.

**FIGURE 3A-C.**
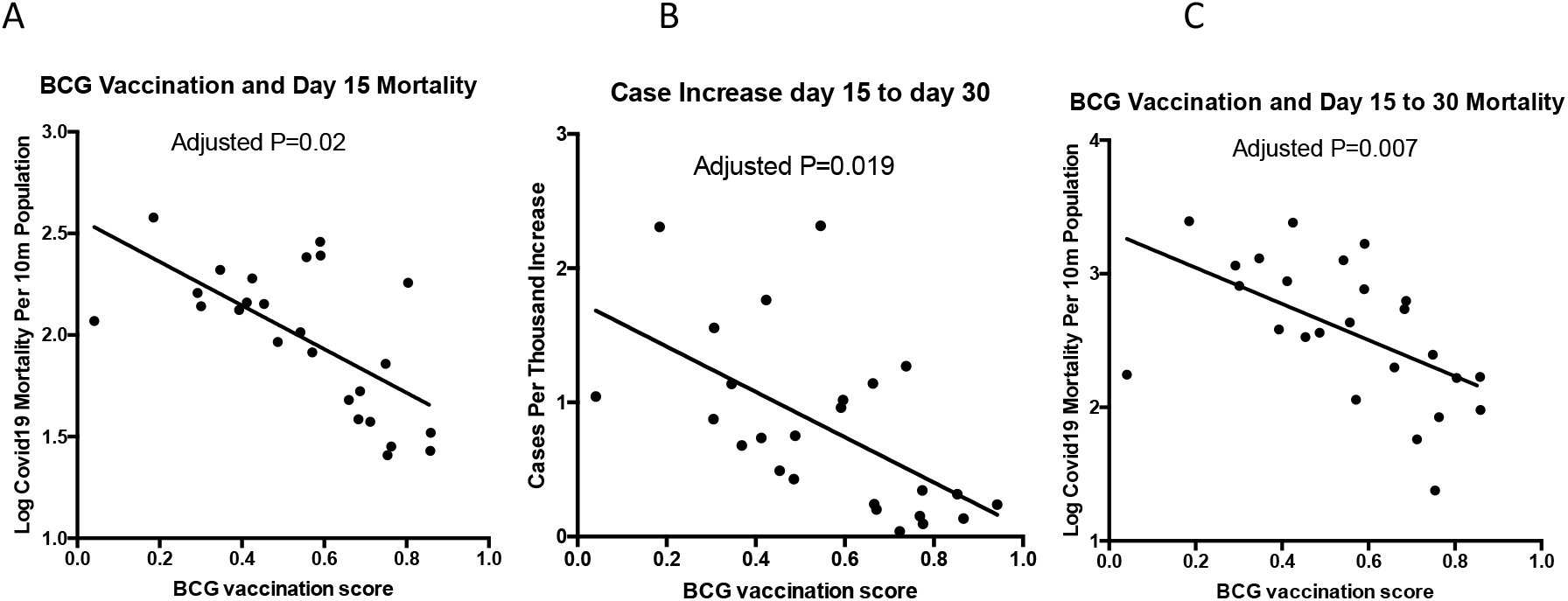
Secondary outcome data for the 25 countries included in the analysis between BCG vaccination score and the following: day 15 mortality per 10 million population (A), univariate P = 0.0009; increase in cases between day 15 and day 30 (B), univariate P = 0.002; increase in mortality per 10 million population between day 15 and day 30 (C), univariate P = 0.0065. The individual figures show P values following adjustment for population size, median age, population density, the proportion of population living in an urban setting, life-expectancy, the elderly dependency ratio (or proportion over 65 years), net migration, days from day 1 to lockdown and case-fatality rate. Abbreviations: BCG, Bacille Calmette-Guérin; PNEUMOCOCCAL, pneumococcal; Influenza, influenza.

## Discussion

This observational, ecological study amongst a series of 25 level 4 European countries suggests that previously reported associations between BCG vaccination and reduced mortality from COVID 19 remain after adjusting for a number of important confounding variables. Similarly, adjusted inverse associations between BCG vaccination and increase in cases and increase in deaths from day 15 to day 30 in the time-course of the outbreak were observed. The covariates used to adjust the relationship included population size, median age, population density, the proportion of population living in an urban setting, life-expectancy, the elderly dependency ratio (or proportion over 65 years), net migration, days from day 1 to lockdown and case-fatality rate. Conversely, associations were not observed with most recently available adult pneumococcal vaccine coverage or seasonal influenza vaccination in this setting with similar covariate adjustment. Prospective clinical studies on the potential role of BCG vaccination repurposed in the setting of Covid 19 will provide important and timely information for clinicians, patients and healthcare systems.

Studies suggesting that countries providing universal BCG vaccine coverage have decreased mortality with COVID 19(6–9) have been the source of wide media interest. However, there have been significant concerns about confounding factors and the lack of randomised controlled data to determine if there is a true causation between BCG vaccination and reduced mortality with COVID 19 (10). While this ecological study may be the first to evaluate BCG vaccination in models with standardisation of mortality rates for timeframe of the outbreak as well as population size, as well as incorporating extensive covariate adjustment related to population structure, the major limitation is that it demonstrates correlation but cannot definitively assign causation. There may be many other confounding variables, which could account for the relationship between BCG vaccination and mortality. Furthermore, it is well accepted that differences in recording mortality between countries have been reported which may affect numbers. Finally, the BCG vaccination score is an estimate of the population coverage with BCG and may not accurately reflect the true population vaccination.

Conversely, the extensive covariate adjustment as well as differential relationship between BCG and influenza/pneumococcal vaccination scores, may support the hypothesis that BCG vaccination is associated with protection from the severest consequences of Covid 19. There is a persistent relationship between BCG vaccination and a variety of mortality and morbidity measures, including peak standardised Z-scores reported for the pandemic. The present study builds on a growing body of evidence that BCG vaccination may be associated with decreased illness severity unrelated to mycobacterium infection. Many have been epidemiological studies looking at overall mortality rates but others have looked at specific unrelated causes such as parasites(11), fungi(12) and bacteria(13) in humans and animals. A recent systematic review concluded that there is evidence with low to moderate risk of bias that BCG vaccination prevents respiratory infections (pneumonia and influenza) in children and the elderly(14) A number of trials are ongoing at present to elucidate the role of BCG in preventing severe COVID 19 with three reported in http://clinicaltrials.gov(15–17).

To date, the nature of non-specific effects with BCG has not been fully elucidated, yet there are a number of possible mechanisms of causation to explain the correlations described in our study. Vaccines with non-specific effects would induce reprogramming of the innate immune responses, a mechanism that clearly differs from the adaptive immunity induced by the antigen-specific responses to the vaccine. Also, since BCG is generally administered to children, any putative innate immune cell “priming” following BCG would, necessarily, be retained many years following vaccination, suggesting an epigenetic link. Interestingly, BCG vaccination has been shown to cause epigenetic changes in innate immune cells via histone-3-lysine-4 (H3K4) methylation [13], which can can augment human IFN and ISG responses to non-mycobacterium and even fungal infections. Therefore, the BCG “primed” innate immune system could counter the SARS-like coronavirus early IFN and ISG suppression, providing a more robust initial innate immune response to limit replication of SARS-CoV-2virus.[18]

Interestingly, a major distinguishing feature of SARS-like coronavirus infections (SARS-Cov-1, MERS and SARS-Cov-2) is their association with innate immune system dysregulation, especially of early host interferon (IFN) and interferon-sensing-gene (ISG) immune responses.[19,20] The early IFN repression in innate immune cells by SARS-Cov1 is in contrast to non-SARS coronaviruses (e.g. 229E and HKU1), which are responsible for milder, cold-like infections.[21] Recently, it was shown that early suppression of IFN and ISG responses by SARS-Cov1 and MERS in human epithelial cells is associated with epigenetic changes involving histone lysine methylation, as has been demonstrated.[20] Furthermore, epigenetic modification of H3K27 in human cells modulates ACE2 expression, the human receptor for SARS-like coronaviruses[22]. The genetic homology of SARSCov1 and SARS-Cov2, indicates that these epigenetic modifications of histone-3-lysine-27 may also feature in Covid 19.

The search for a vaccine to manage the SARS-CoV-2pandemic is a global healthcare priority because of high transmissibility and Covid19 mortality rates. However, following a small, non-human primate study of the Oxford RNA vaccine, the world’s largest producer of vaccines is in negotiation to dedicate its production capacity to this developmental vaccine for SARS-CoV-2. [23,24] However, this could limit normal childhood vaccination programmes despite risks of using novel technology platforms to treat the novel SARS-CoV-2 virus. While we await the results of prospective, randomised, controlled clinical studies on novel vaccines specific for SARS-CoV-2, recent clinical progress with known drugs such as remdesivir highlight the potential of repurposing known therapies to combat Covid-19(18). The present study suggests that ongoing, prospective clinical studies to determine if BCG vaccination reduces the severity of Covid-19 in vulnerable patients could provide important information on the potential for innate immune priming with BCG as an adjunct to other clinical strategies for protection against the worst consequences of SARS-Cov2.

## Conclusion

The association of BCG and reduced mortality from Covid-19 was seen even after standardising mortality between level 4 European countries and adjusting for confounding variables relating to population structure. A similar association was not seen with adult influenza and pneumococcal vaccination. The results of prospective clinical studies on BCG vaccination as an adjunct to usual care in the setting of Covid-19 are awaited.

## Data Availability

Publicly available data used and referenced

